# Introduction of Dengue Virus Serotype 3 in the Afar Region, Ethiopia

**DOI:** 10.1101/2024.07.19.24310689

**Authors:** Feleke Mekonnen, Bilal A. Khan, Endalkachew Nibret, Abaineh Munshea, Daniel Tsega, Demeke Endalamaw, Senait Tadesse, Gizachew Yismaw, Damtie Lankir, Jemal Ali, Mariana Ulinici, Emanuele Orsini, Ursa Susnjar, Tea Carletti, Danilo Licastro, Molalegne Bitew, Marta Giovanetti, Alessandro Marcello

## Abstract

The genetic analysis of the Dengue virus circulating in Ethiopia’s Afar region, in 2023, identified three distinct introductions with spatiotemporal clustering linked to genomes from Asia and Italy. These findings are crucial for enhancing prevention and control strategies, reinforcing the necessity to provide sustainable tools for genomic epidemiology in Africa.

## Introduction

Ethiopia has been experiencing annual dengue outbreaks since the first cases were reported in Dire Dawa in 2013 (1). Subsequent outbreaks occurred in the Eastern and Southern districts, including Godey town and Arbaminch, in 2020 (2). In 2019 an outbreak occurred in the Gawane District of the Afar Region in Noth-Eastern Ethiopia (3). Dengue serotypes 1 and 2 (DENV-1 and DENV-2) have been reported in the country (4,5), while dengue virus serotype 3 (DENV-3) was detected only recently in the Afar Region (6). Despite the increasing number of cases, much is still unknown about the genomic diversity and evolution of DENV lineages currently circulating in the country. To address this gap, a cross-sectional study was conducted from June to October 2023 on 384 acute febrile (AFI) patients attending three healthcare facilities in the Afar Region: Logia, Mille and Gewane. The study was conducted within EXPANDIA (Expanding Diagnostics and Surveillance in Africa), a project implemented by the International Centre for Genetic Engineering and Biotechnology (ICGEB).

### The Study

Patients presenting at Logia, Mille, and Gewane healthcare facilities in the Afar Region exhibiting symptoms typical of dengue were included in this study. These symptoms comprised an acute onset of high fever persisting for 2-7 days, severe joint and muscle pain, headache, retro-orbital pain, rash, nausea/vomiting, and unusual bleeding, or bruising. We included 384 patients who visited these facilities between June and October 2023. The Regional Public Health Research Ethics Review Committee, Ethiopia, approved the study (Ref. NoH/R/T/T/D/07/39). Sociodemographic and clinical information, along with other relevant data, was gathered using a structured questionnaire (**Appendix 1 Table 1&2)**.

Malaria infection was excluded using a smear test on whole blood. Serum was obtained from venous puncture and tested on-site using the Wondfo One Step Dengue IgG/IgM test (Guangzhou Wondfo Biotech Co., Ltd., China). Serum samples that tested positive for DENV-specific IgM (32.8%, 126/384) and/or IgG (36.2%, 139/384) were transported at -20 °C to Addis Ababa (APHI and BETiN) and to ICGEB. Viral RNA purification from serum was performed using the QIAamp Viral RNA Mini Kit (Qiagen, Germany), and DENV RNA was detected using the Luna Universal Probe One-Step RT-qPCR Kit (NEB) using in-house pan-Dengue primers and probe. Positive samples were assigned to the DENV-3 serotype by amplifying the C-prM region, as previously described (7), followed by Sanger sequencing and data analysis using the Genome Detective DENV typing tool (8). Samples (n=7) with a cycle threshold (Ct) value of ≤32 and available epidemiological metadata - such as the date of symptom onset, date of sample collection, sex, age, municipality of residence, symptoms, and disease classification, were subjected to a nanopore whole genome sequencing using a set of tiled primers (9) following previously described protocols with slight modifications (10). For amplification of DENV-3 genome, 30 primers were used (7) to cover the almost entire coding sequence (CDS) of the DENV-3 genome (positions 132 to 10139) in 800-900 bp amplicons (**Appendix 2 primer list & protocol**). We reconstructed phylogenetic trees to explore relationships between sequenced genomes from Ethiopia (n=7) and other global regions (n=351) (**Appendix 3**).

The sequencing procedure yielded an average coverage of 89%, ranging from 73.8% to 96.9% (**Figure 1a**). This allowed for the identification of the DENV-3, genotype III, major lineage B, following the recent release of the DENV classification system (11). Genome sequences were obtained from all three districts of the Afar region: Mille, Logia, and Gewane (**Figure 1b**).

**Figure 1.**
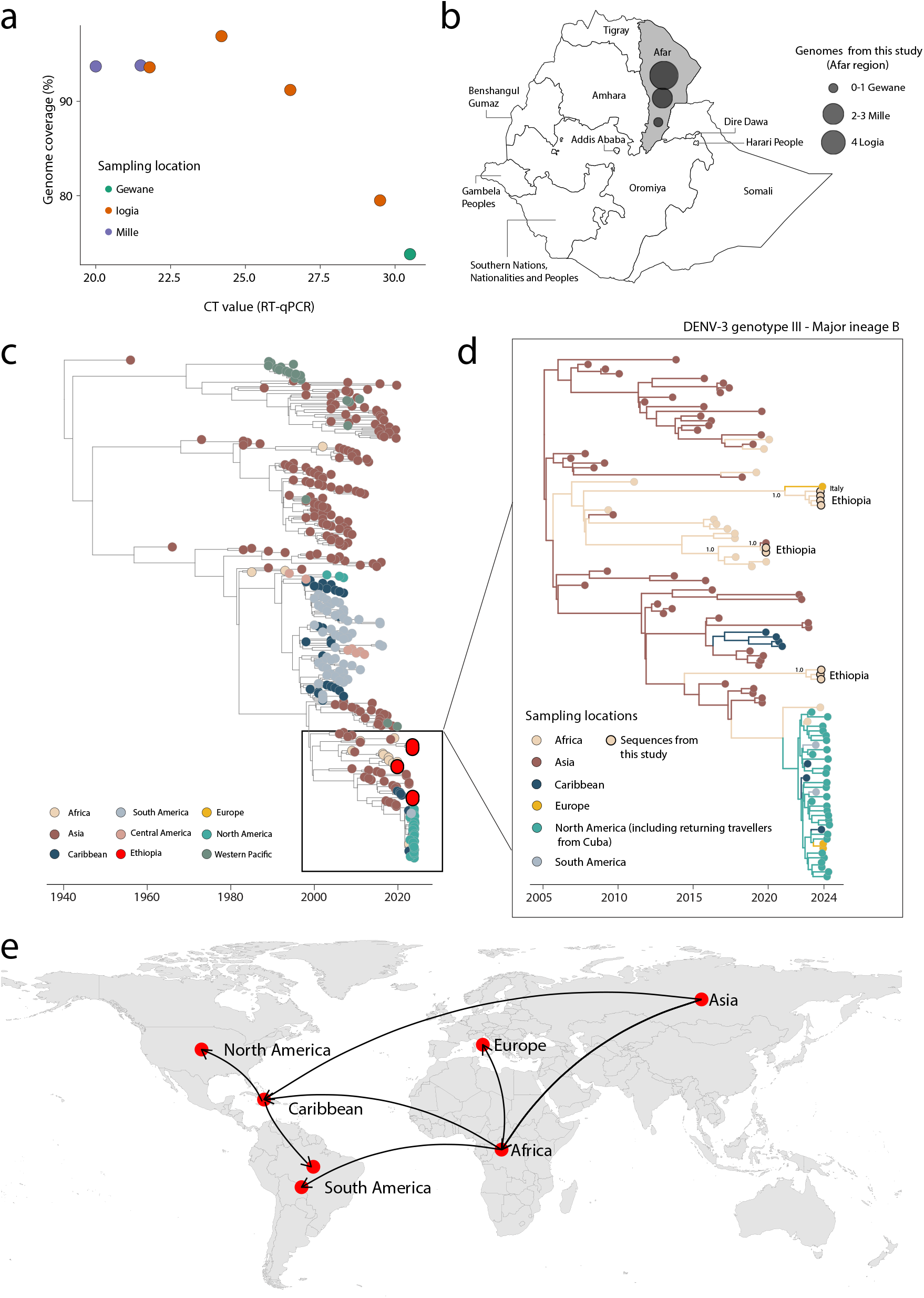
Dynamics of DENV-3 in Ethiopia. a) The percentage of DENV genome sequenced plotted against RT–qPCR Ct value for each sample (n = 7). Each circle represents a sequence recovered from an infected individual in Ethiopia and is coloured by sampling location; b) Map of Ethiopia showing the sampling locations of new DENV-3 genome sequences by region and district. The size of the circles represents the number of new genomes generated in each district; c) Time-stamped Maximum Likelihood tree including the novel DENV-3 sequences (n=7) generated in this study, along with n=351 reference strains belonging to DENV-3 genotype III sampled worldwide. Tips on the tree are annotated with circles and colored based on their locations; d) The inset tree on the right represent a Maximum Clade Credibility tree inferred using a smaller dataset (n=111) which included all novel genomes from Ethiopia. Each branch is colored according to the legend on the left; Ethiopian sequences are marked with a black circle. Support for branching structure is shown by posterior probability values at key nodes; e) Representation of the spatiotemporal reconstruction of the spread of DENV3-III Major lineage B.

To understand the phylogenetic history of DENV-3, genotype III, we combined our recently sequenced samples (n=7) from 2023 with a global set of reference samples. Our analysis revealed that the new genomes cluster together within the major lineage B and that three independent introduction events have occurred in Ethiopia over time (**Figure 1c**).

We then investigated the spatial-temporal dynamics of the Ethiopian DENV-3, genotype III, in more detail using a smaller dataset (n=111) derived from the major lineage B, which has an Asian origin (**Figure 1d**). Since 2015, this lineage has spread multiple times into Africa, the Caribbean, and subsequently to North and South America, where it was recently described as a re-emerging variant circulating in the region (**Figure 1e**) (12). Within the Ethiopian sequences, we identified three distinct clades. Clade I dates back to early February 2021 (95% Highest Posterior Density -HPD: April 2019 to February 2022) and includes Ethiopian strains that cluster monophyletically with a genome sequence isolated in Rome, Italy, in 2023. Although genetic data alone cannot provide evidence of transmission direction and potential cryptic transmission must be considered due to the paucity of genomes from other locations, our analyses indicate strong clustering support for the Ethiopian sequence with the one recovered in Italy (posterior probability support = 1.0). Clade II, dating back to late June 2019 (95% HPD: January to October 2019), comprises two newly identified Ethiopian strains that cluster with a strain isolated in Asia, suggesting a potential introduction from Asia to Ethiopia. Clade III, dating back to July 2022 (95% HPD: April 2021 to October 2022), consists solely of sequences from Ethiopia and shows a clear Asian origin. Given the limited sequences from other locations and the low number of genomes from Ethiopia, this clade might represent persistent local circulation within the country. Together these findings highlight the dynamic nature of DENV-3 transmission and the complex epidemiological patterns in the country.

## Conclusions

This study underscores the dynamic and complex nature of DENV-3 transmission within Ethiopia. The introduction of at least three independent DENV-3 lineages into the Afar region, as indicated by genomic evidence, highlights a significant epidemiological pattern. These introductions have connections with distinct global regions including Asia and Europe, particularly Italy, suggesting international travel and movement as potential vectors for disease spread. Temporal and phylogenetic analyses reveal that while some of the identified strains suggest recent introductions, others indicate more established, ongoing local transmission within the country.

These findings are critical for the Ethiopian public health response, emphasizing the need for robust surveillance systems that incorporate genomic epidemiology to track virus evolution and spread effectively. Such efforts are essential not only for timely and targeted dengue control measures but also for preparing for potential outbreaks of other imported viral diseases. This study also reinforces the importance of international collaboration in genomic surveillance to better understand and mitigate the spread of infectious diseases globally.

Additionally, this study produced near-complete genomes from the Afar region for the first-time using Oxford Nanopore sequencing technology. This approach could be implemented in Ethiopia and other African countries to enhance genomic surveillance and management of DENV transmission in the region.

**Table 1:** Sociodemographic Characteristics of Dengue cases in Afar Region, Ethiopia. June-October, 2023.

| Socioeconomic Status of AFI cases |  |  |  |  |  |
| --- | --- | --- | --- | --- | --- |
| Variables |  | Dengue IgM |  | COR (95%CI) | P-value |
|  |  | Positive N (%) | Negative N (%) |  |  |
| Gender |  |  |  |  |  |
|  | Male | 102(32.4) | 213(67.6) | 1(0.643_1.928) | 0.7 |
|  | Female | 24(34.8) | 45(65.2) | 0 |  |
| Age(yr) |  |  |  |  |  |
|  | (6-15) | 3(2.4) | 7(2.7) | 0 |  |
|  | (16-45) | 50(39.7) | 103(39.9) | 0.85(0.214-3.388) | 0.82 |
|  | (>46) | 73(57.9) | 148(57.4) | 0.91(0.226-3.668) | 1 |
| Residence |  |  |  |  |  |
|  | Urban | 56(37.7) | 101(64.3) | 1.2(0.745-1.972) | 0.44 |
|  | Semi-Urban | 27(30) | 63(70) | 1.3(0.741-2.251) | 0.37 |
|  | Rural | 43(31.4) | 94(68.6) | 0 |  |
| Marital Status |  |  |  |  |  |
|  | Single | 84(31.5) | 183(68.5) | 0.8(0.519-1.295) | 0.39 |
|  | Married | 42(35.9) | 75(64.1) | 0 |  |
| Occupation |  |  |  |  |  |
|  | Governmental | 3(37.5) | 5(62.5) | 0 |  |
|  | Pastoralist | 57(32.8) | 117(67.2) | 1.2(0.284-5.335) | 0.78 |
|  | Unemployed | 34(39.5) | 52(60.5) | 1.2(0.262-5.836) | 0.79 |
|  | House wife | 16(32.7) | 33(67.3) | 0.92(0.206-4.093) | 0.91 |
|  | Merchant | 3(33.3) | 6(66.7) | 1.2(0.164-8.799) | 0.86 |
|  | Others | 13(22.4) | 45(77.6) | 2(0.437-9.871) | 0.36 |
| Health Facility |  |  |  |  |  |
|  | Logia health center | 89(40.6) | 130(59.4) | 2.2(0.963-5.139) | 0.61 |
|  | Mille health center | 29(22.1) | 102(77.9) | 0.9(0.378-2.258) | 0.86 |
|  | Gewane health center | 8(23.5) | 26(76.5) |  |  |

0 = reference group, \* = statistically significant ( $p\text{-value} < 0.05$ ), COR (95%CI) = crudes odds ratio at 95% confidence interval

**Table 2:**
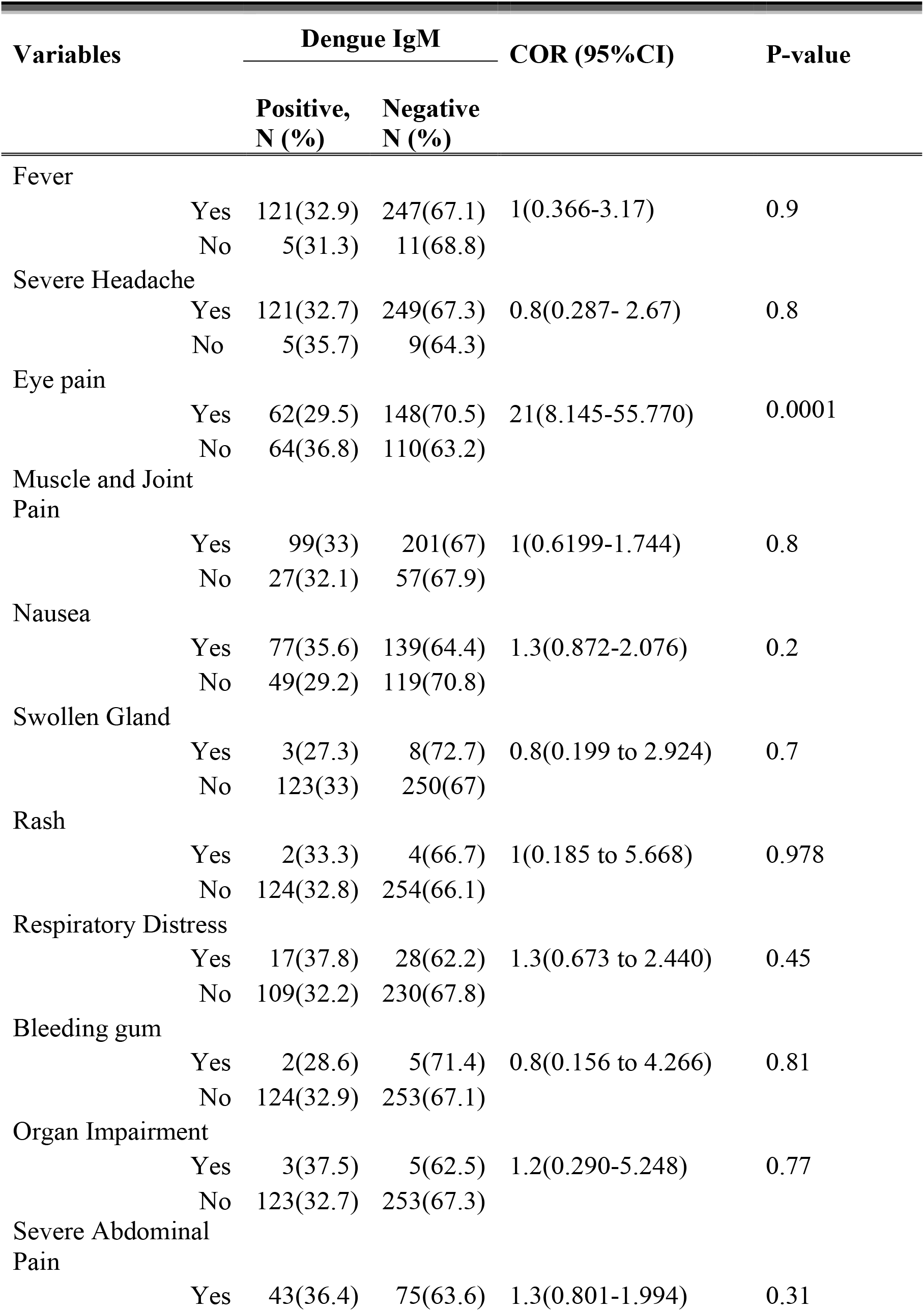
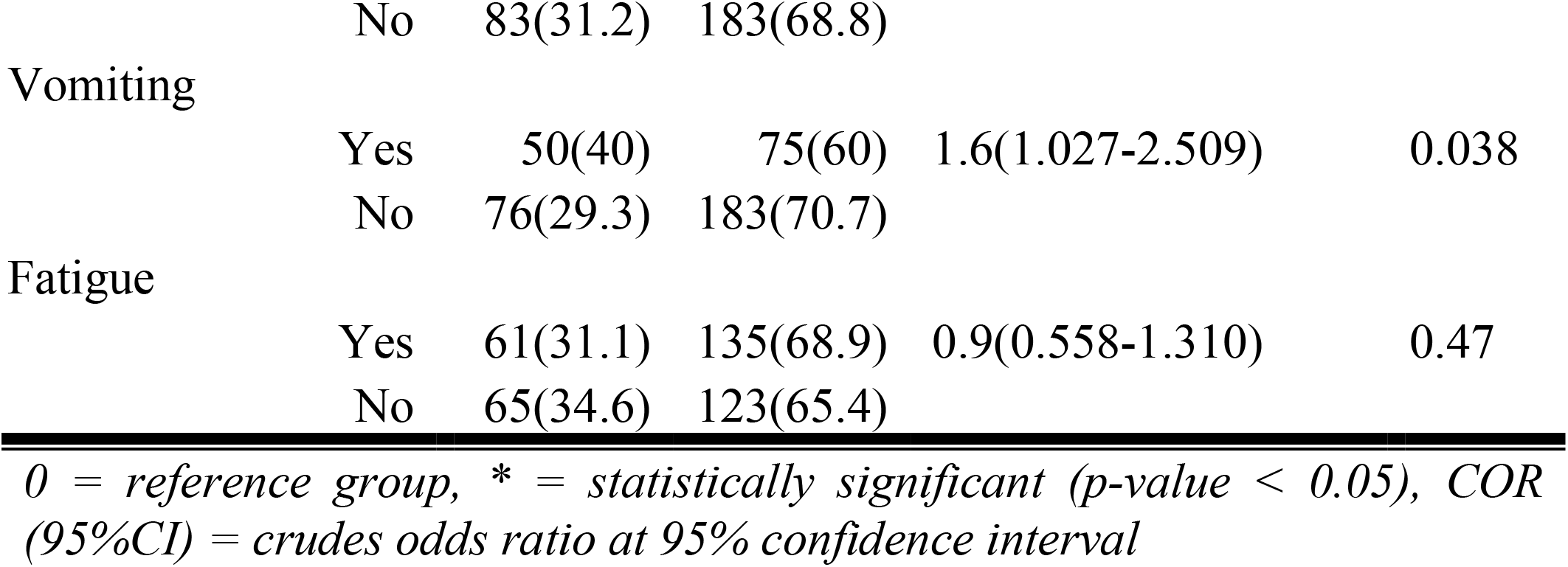
Clinical Characteristics of Dengue patients in Afar region, Ethiopia, June-October 2023.

**Table 3:**
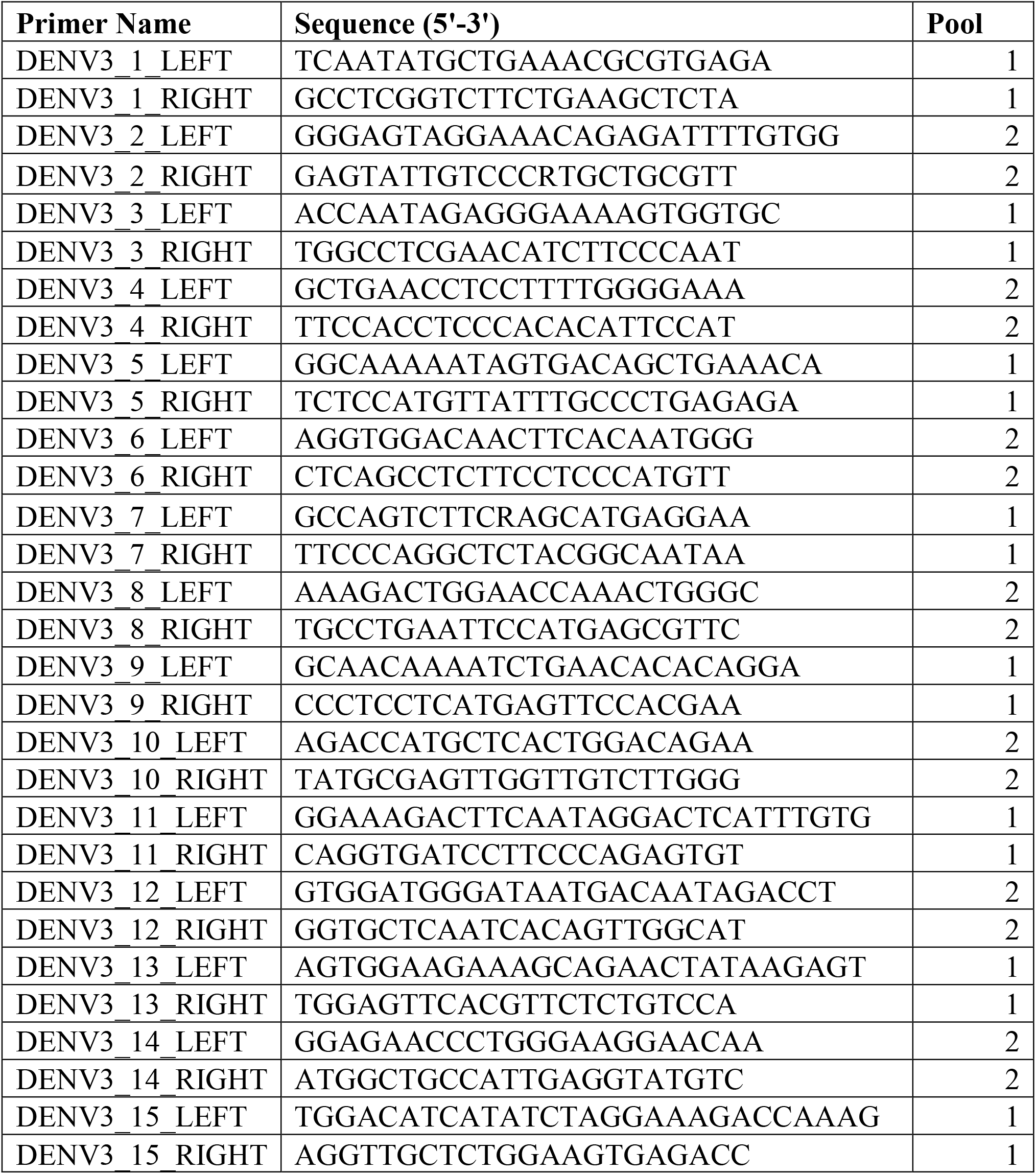
Primers used for DENV-3 Sequencing.

## Data Availability

All data produced in the present work are contained in the manuscript.

This work was supported, in whole or in part, by the Bill & Melinda Gates Foundation [Grants No. INV-034,799 and INV-064464]. Under the grant conditions of the Foundation, a Creative Commons Attribution 4.0 Generic License has already been assigned to the Author Accepted Manuscript version that might arise from this submission. Other sources of funding include the ICGEB CRP to Ethiopia (CRP/ETH21-01) and PNRR “GENESIS” - COC-1-2023-UNIPV - ID S1.P0002 (INF-ACT).

The content and findings reported in this work are the sole responsibility of the researcher(s) involved and do not necessarily reflect the official positions or sentiments of the funding agencies that supported this project.

## About the Author

Feleke Mekonnen is a PhD student at Bahir Dar University, Ethiopia. His research interests include viral and bacterial infections. Currently, he is a visiting researcher at the Molecular Virology Laboratory, ICGEB, Trieste, Italy.

## PROTOCOL: Brief ONT sequencing protocol for DENV-3^**1**^

Briefly 12 μl of RNA was reversely transcribed using 3 μl of Luna Script RT Super-Mix Kit (New England Biolabs, U.S.) at a thermocycler following temperature cycles of 25 °C for 2 min, 55 °C for 10 min, and 95 °C for 1 min, followed by DENV3-specific multiplex PCR in 2 pools by using Q5 Hot Start High-Fidelity 2x Master Mix (New England Biolabs, U.S.) with the cycling conditions of 98 °C for 30 seconds followed by 40 cycles of 95 °C for 15 seconds, 50 °C for 30 seconds, 72 °C for 60 seconds & final extension of 72 °C for 2 minutes. The resulting PCR products were quantified by the Qubit dsDNA High Sensitivity assay on a Qubit 4 fluorometer (Thermo Fisher Scientific, USA). The products were pooled in 1:1 ratio and purified by SPRI PCR Clean Dx beads (Aline biosciences, U.S.) which were further subjected to end-preparation by NEBNext® Ultra™ II End Repair/dA-Tailing Module (New England Biolabs, U.S.) and barcoded by Native Barcode Expansion Kit-NBD196 (Oxford Nanopore Technologies, UK). The barcoded samples were consolidated and cleaned by SPRI beads as described above followed by AMII adaptor ligation by NEBNext® Quick Ligation Module (New England Biolabs, U.S.). The final library was SPRI cleaned up, quantified and loaded into R9.4.1 flow Cell using the Ligation Sequencning Kit-LSK110 (Oxford Nanopore Technologies, UK). The sequencing data were collected for 16 hours, and the raw reads were base called and de-multiplexed in MinION Mk1C in-built tools in Minknow v 22. The resulting FASTQ files were used to generate consensus sequences by using de novo assembly in Genome Detective web tool (https://www.genomedetective.com/).

GenBank accession numbers for the nucleotide sequence(s):

SUB14602032 DENV_Ethiopia-54 PQ014884

SUB14602032 DENV_Ethiopia-51 PQ014885

SUB14602032 DENV_Ethiopia-52 PQ014886

SUB14602032 DENV_Ethiopia-53 PQ014887

SUB14602032 DENV_Ethiopia-55 PQ014888

SUB14602032 DENV_Ethiopia-49 PQ014889

SUB14602032 DENV_Ethiopia-58 PQ014890

### Phylogenetic and Phylogeographic Analysis Method

Sequence alignment was done using MAFFT and edited with AliView. A preliminary phylogenetic tree was built using IQ-TREE 2 with the GTR+G4 model (13) signal was evaluated with TempEst, and outliers were removed before creating a time-scaled tree using TreeTime. Phylogenetic relationships were inferred using a smaller dataset which included the Ethiopian strains (n=111) using BEAST v1.10.4 with a relaxed lognormal clock, skygrid population size, and GTR model. Phylogeographic analysis considered six locations: Asia, Africa, North America, Europe, South America, and the Caribbean. The analysis used an empirical distribution of 1000 trees, running the MCMC chain for 100 million iterations, sampling every 1000. Maximum clade credibility (MCC) trees were summarized using TreeAnnotator v1.10.4 and visualized with the ggtree package in R. Geographic visualization was performed using SpreaD3.

## Footnotes

1 A detailed version of this protocol is available on request.

